# Acute and chronic effects of local muscle vibration training inducing illusions on wrist strength and neurophysiological measures

**DOI:** 10.1101/2025.04.24.25326331

**Authors:** Sophie Julliand, Jérémie Gaveau, Alain Martin, Nicolas Amiez, Adrien Guzzo, Martine Lemesle-Martin, Davy Laroche, Charalambos Papaxanthis

## Abstract

Local muscle vibrations (LMV) acutely and chronically influence the sensorimotor system, but their effects depend on application conditions. The mechanisms underlying repeated LMV exposure remain debated. This study examined the chronic effects of LMV under illusion-inducing conditions on the upper limb.

Nineteen healthy participants underwent 9 LMV sessions (80 Hz, 20 min) over 11 days targeting the right wrist flexors. Illusions were assessed using subjective scales and EEG (alpha band desynchronization). Neurophysiological parameters (spinal, corticospinal excitabilities, and intracortical inhibition) and grip strength were evaluated before and after a control session (20-min rest) and an LMV session, as well as after 9 LMV sessions and 5 days post-protocol.

Participants consistently experienced illusions throughout the protocol, with stronger perceptions in the first 15 minutes. Yet, subjective and objective measures were found independent. Acute LMV significantly reduced the H-reflex (-45 ± 38%) but did not alter strength or other neurophysiological measures. More, repeated LMV exposure induced no chronic changes in strength or neural excitabilities/inhibition.

While confirming acute LMV effects under illusion conditions, this study found no evidence of chronic adaptations. It also suggests that subjective and objective (EEG-based) illusion measures reflect distinct neurophysiological mechanisms.

## Introduction

Local Muscle Vibrations (LMV) are a non-invasive neuromodulation technique that strongly stimulates local sensory receptors. Since the 1970s, LMV has been widely used to explore the sensorimotor system^1–4^. More recently, their potential therapeutic applications have gained attention, particularly in neurological rehabilitation^5,6^. Despite this growing interest, the mechanisms underlying the LMV-induced modulations remain incompletely understood, particularly in the context of repeated applications.

In healthy participants, acute application of LMV (a single session) has been demonstrated to produce temporary neuromuscular modifications in the vibrated site, mainly including transient changes in muscle strength, intrinsic motoneuronal excitability, and a significant reduction in spinal excitability as reflected by a decrease in Hoffmann’s reflex (H-reflex)^4,7–10^. In addition to spinal adaptations, acute enhancement of cortical excitability has been suggested following LMV. Evidence from neurophysiological studies, which compared motor evoked potentials (MEPs) with cervicomedullary (CMEPs) or thoracic motor evoked potentials (TMEPs), has demonstrated increased MEP/CMEP or MEP/TMEP ratios following LMV^7,11,12^. While MEP reflects corticospinal excitability, CMEP and TMEP serve as indicators of motoneuronal excitability. Findings from electroencephalography (EEG) further support these observations, as they revealed strong desynchronization in the alpha power spectrum over the contralateral sensorimotor cortex (electrodes C3 or C4), indicating increased cortical activity during LMV that may persist post-vibration ^13,14^. While some research partly attributes these findings to intracortical excitability changes – such as reduced intracortical inhibition and increased facilitation^15,16^– the results remain inconsistent, with other studies reporting no significant effects on these parameters^17^.

The variability of LMV effects likely stems from differences in application parameters^4^, muscle state (i.e., contracted or relaxed), LMV parameters (i.e., frequency, amplitude, duration of the stimulation), and contextual factors (i.e., visible or hidden limb)^10,18–20^. A particularly relevant distinction is whether LMV induces movement illusions. For instance, when LMV is carefully applied on a relaxed and hidden limb, with frequencies ranging from 70 to 110 Hz and small amplitude (< 3mm)^20,21^, the literature consensually reports that LMV can create the sensation of movement illusions, giving the impression that the vibrated muscle is being stretched^22^. These illusions arise from the heightened activation of neuromuscular spindles by LMV, which generate robust proprioceptive signals via Ia afferent fibers that project towards the spinal cord up to the brain^23^. Research has shown that LMV-inducing movement illusions elicit greater cortical excitability, compared to LMV without illusions. For instance, functional MRI studies have demonstrated significant activation of the premotor, sensorimotor, and parietal cortices during LMV, with a larger activation when movement illusions are present^24,25^. Interestingly, these cortical regions overlap with those activated during real movements^26^, suggesting that movement illusions engage comparable neural structures as real movements, albeit to a lesser extent. EEG studies have reported corroborating findings, showing a strong modification of activity in the alpha or Mu band (8-13 Hz) during LMV, which becomes even more pronounced when movement illusions occur^27,28^. Recent work by Amiez et al. (2024a)^18^ compared two LMV conditions both applied to a relaxed limb; one inducing movement illusions and the other without illusion but with Tonic Vibratory Reflex (TVR). Their findings revealed distinct acute after-effects on muscle strength; specifically, the vibrated muscle experienced a greater strength reduction in the TVR condition. These results highlight the importance of precisely controlling LMV parameters, as the presence or absence of movement illusions can differentially modulate neurophysiological and functional responses following LMV.

While LMV is increasingly used in clinical rehabilitation, often applied multiple times per week over several weeks^29,30^, studies examining its chronic effects (repeated applications) remain limited. Such research is critical to understanding the underlying neural mechanisms and optimizing their application. Existing studies suggest that short- and long-term adaptations of repeated LMV may differ^4,19,31–34^, further emphasizing the need for controlled investigations. Notably, the effect of LMV- induced movement illusions in repeated sessions has not been systematically explored. To address this gap, our study investigated both the acute (following a single session) and chronic (following repeated sessions) effects of LMV-inducing illusions on wrist flexors. We assessed movement illusions using subjective scales and EEG measures. Key neurophysiological outcomes included corticospinal excitability (MEPs), spinal excitability (H-reflex), intracortical inhibition (SICI), and functional performance (maximal grip strength). These parameters were measured at multiple time points throughout the LMV protocol. We hypothesized that, acutely, our protocol would replicate the recent findings showing no significant modifications of strength when LMV were applied under illusion-inducing conditions^18^. Additionally, we expected distinct chronic effects on grip strength (i.e., no change) compared to previous LMV training studies^33,34^, as the presence of illusions may influence this parameter differently. Lastly, we aimed to identify the underlying neurophysiological adaptations associated with this specific LMV protocol.

## Methods

### Participants

Nineteen (n = 19) volunteers were included in this protocol (9 females and 10 males, mean age 26.6 ± 5.3 years). All participants were right-handed (mean score 0.8 ± 0.2) as determined by the Edinburgh Handedness Inventory^35^. None of the volunteers had any neurological, psychological, or musculoskeletal diseases. The protocol was approved by CPP Ile de France IV (number 2021-A03219- 32, and ClinicalTrials NCT05315726), and all participants provided verbal informed consent, aligning with the revised 2013 Declaration of Helsinki.

### General experimental setup

In visit 1 (V1) and visit 2 (V2), we scheduled two blocks of measures (PRE and POST, see Figure 1), separated by a rest of 20 minutes between V1 PRE (V1_PRE_) and V1 POST (V1_POST_), and a 20-minute LMV of the wrist flexors between V2 PRE (V2_PRE_) and V2 POST (V2_POST_). Eleven days separated V1 and V2, a period during which participants were instructed to maintain their normal lifestyle. V1_PRE_, V1_POST,_ and V2_PRE_ were used to assess data reproducibility over time and served as control conditions.

**Figure 1:**
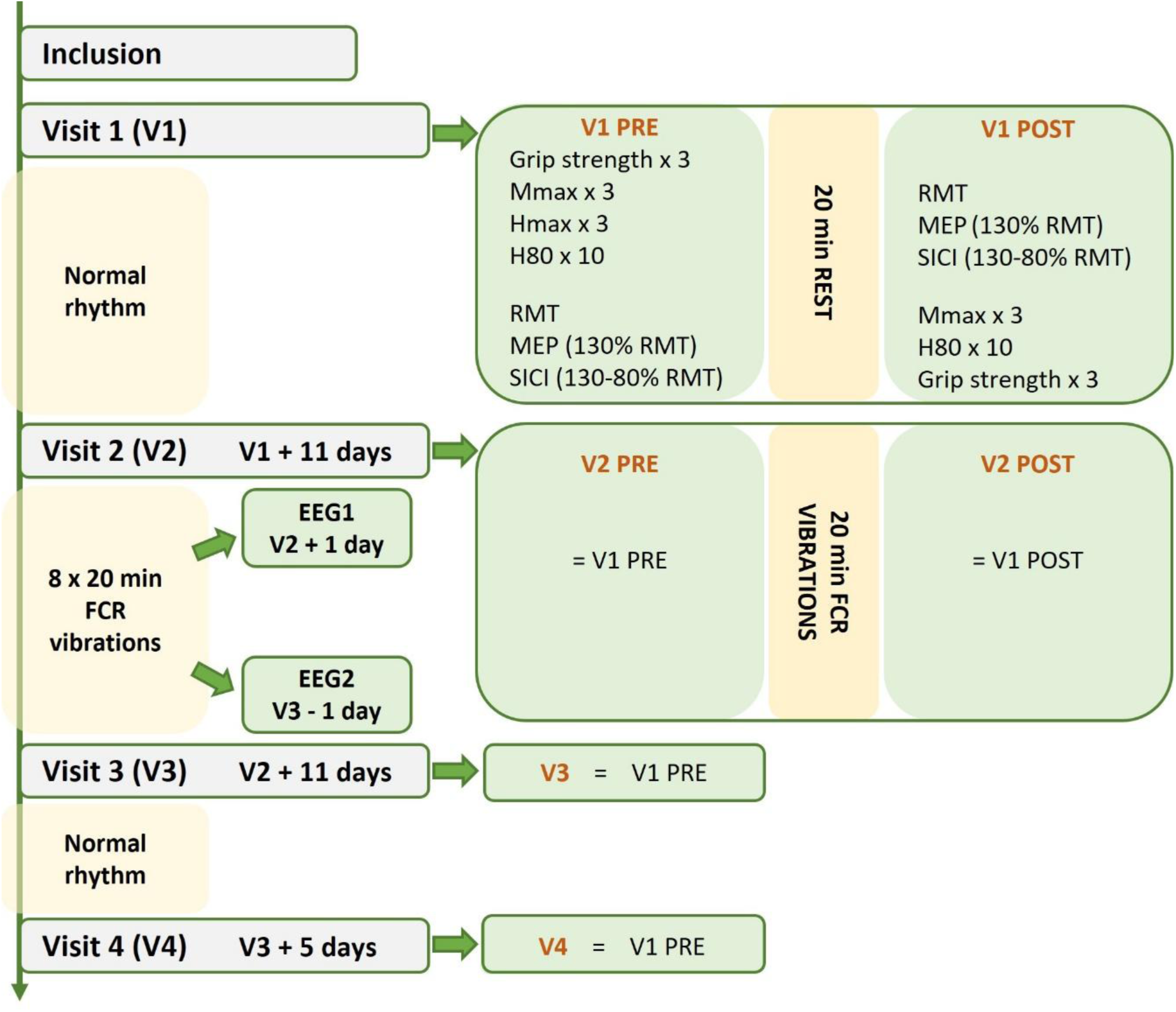
Study design. Evaluations were conducted on the relaxed right flexor carpi radialis (FCR) muscle under identical conditions across the four dedicated visits (V1, V2, V3, and V4). POST tests were administered after 20 minutes of rest in V1 and after 20 minutes of wrist flexors LMV in V2. A total of 9 vibratory sessions were administrated, starting with the first session at V2 and ending with the final session the day before V3. The LMV protocol aimed to optimize movement illusions by vibrating the tendon site, using a frequency of 80 Hz, an amplitude of 2mm, and with the hand and forearm hidden. Illusions were quantified using EEG and a subjective scale during the second and last LMV sessions. The acute effects of LMV were evaluated by comparing the PRE and POST sessions in V1 and V2. The chronic effects of LMV were assessed by comparing V2PRE with V3 and V4. Maximal M-wave (Mmax); Maximal Hoffman reflex (Hmax); 80% of Hmax (H80); Resting Motor Threshold (RMT); Motor evoked potential (MEP); Short intracortical inhibition (SICI); Electroencephalogram (EEG).

The LMV protocol began at V2 (after V2_PRE_) and finished the day before V3. During this period, participants received a total of 9 sessions of 20-minute LMV during which subjective ratings of illusions were evaluated. EEG recordings were performed during the second (EEG1) and last LMV session (EEG2). In V3 and V4, we recorded the same physiological measurements as in V1_PRE_ and V2_PRE_. During the 5 days separating V3 from V4, participants were instructed to maintain their normal lifestyle.

In this protocol, the acute effects of LMV on neurophysiological parameters and strength were assessed by comparing V1_PRE_, V1_POST_, V2_PRE,_ and V2_POST_. To evaluate the training-induced impact of repeated LMV, V2_PRE_ was used as the baseline and compared with V3 and V4.

All the experiments were consistently conducted at the same time of day for each participant to avoid possible intra-participant circadian fluctuations^36,37^. During the physiological measurements (Figure 2A), participants were comfortably seated on a chair, with their trunk in the vertical position, and their right forearm in a semi-pronated position on a custom-made support. The elbow was flexed at 90° and the shoulder abducted at 30°, flexed at 15°, and externally rotated at 20°. During the 20- minute rest (V1) and all the LMV sessions (Figure 2B), participants were seated in a semi-reclined position to promote muscle relaxation and enhance the likelihood of experiencing illusions^20^. Their arm was hidden from view and supported at the elbow and wrist by malleable cushions, with the elbow semi-flexed, pronated, and the wrist in a neutral position.

**Figure 2:**
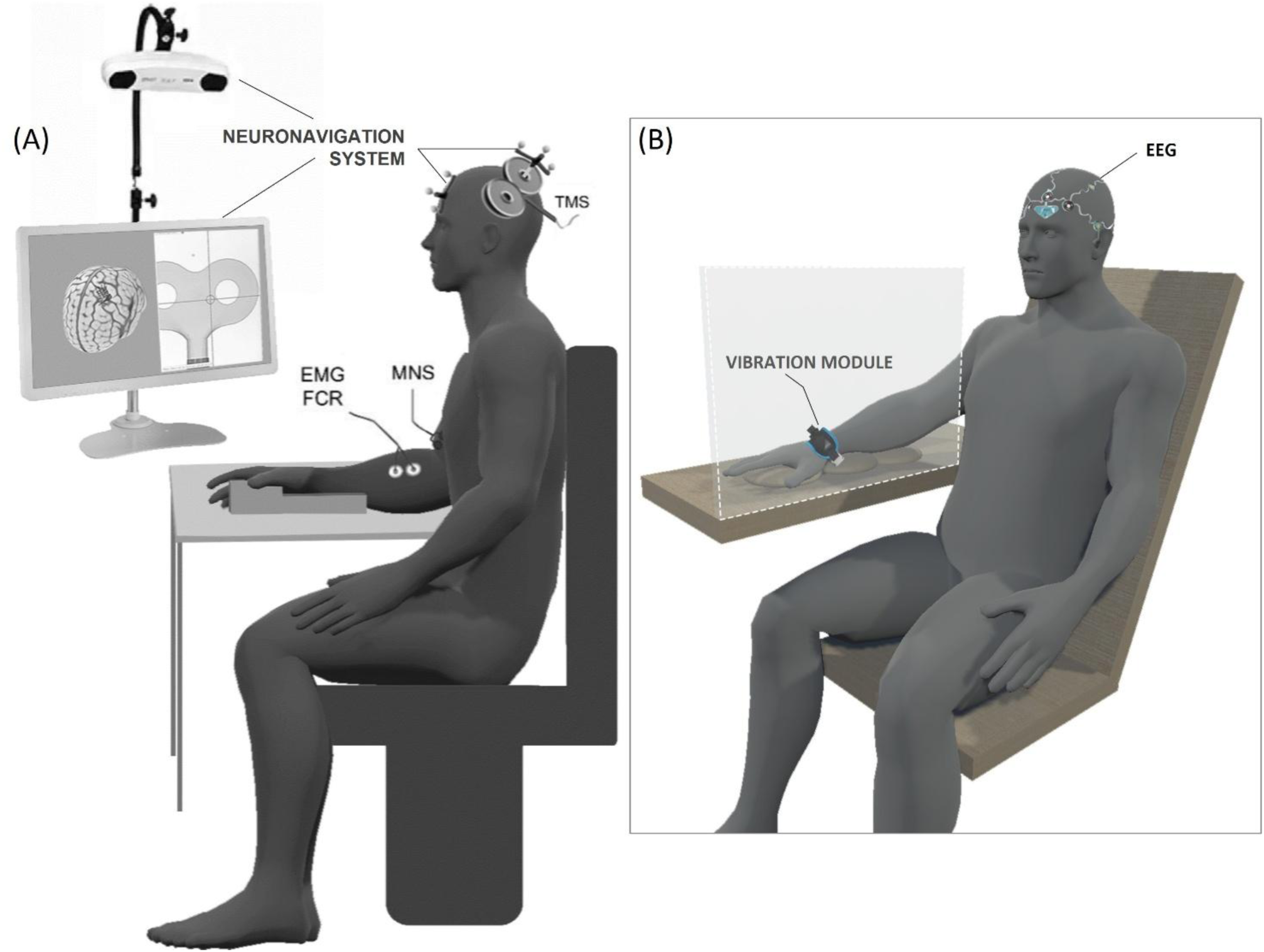
Schematic view of the experimental setup and the participants’ position. Figure 2A shows the measures recorded in the Flexor Carpi Radialis (FCR) with a Neuronavigation system, and with Transcranial Magnetic Stimulation (TMS), Median Nerve Stimulation (MNS), and Electromyography (EMG) devices. Figure 2B illustrates the position of the vibratory device. Note also that an Electroencephalogram (EEG) was performed twice, during the second and last LMV sessions.

### Vibrations

Participants underwent a total of 9 vibratory sessions (1 per day during two consecutive weeks), each consisting of 20 minutes of continuous mechanical LMV. The vibration module (Vibramoov, Technoconcept, Manosque, France) was directly applied to the skin, approximately 2 cm proximal to the right wrist crease, and secured around the forearm with a strap (Figure 2B). To ensure optimal comfort and appropriate pressure, the wristband size was individually adjusted for each participant. Continuous LMV was administered for 20 min at a frequency of 80 Hz with an amplitude of 2 mm (according to the manufacturer’s specifications). The participants’ view of their forearm was obstructed with a cardboard box to induce movement illusions and diminish the tonic vibratory reflex^20,38^. Participants were instructed to relax and concentrate on the LMV. The 20-minute duration was selected based on previous studies demonstrating its effectiveness in inducing movement illusions, and modulating H-reflex and strength^7,10,18^.

### Illusions

During each vibratory session, three visual analog scales were used to rate the participants’ subjective sensations. These scales, ranging from 0 (non-existent) to 10 (very strong), assessed the amplitude of the illusory movement (strength), the duration of the illusory movement during the last minute (continuity), and the clarity of illusions as if a real wrist extension was performed (vividness)^24^. Participants were asked to evaluate their sensation at specific intervals (1 min, 5 min, 10 min, 15 min, and 19 min), providing one score per scale at each time point. The pooled score, calculated as the mean of the three scales, served as the marker for the overall intensity of the illusions.

Two EEGs sessions were conducted during the second and last vibratory session (i.e., EEG1, and EEG2, see Figure 1). EEG signals were recorded (Icecaps, Neuronaute, BioSerenity, Paris, France) from 21 electrodes, based on the 10-20 system. Electrode impedances were kept below 5 kΩ, and signals were recorded with a sampling rate of 500 Hz. The entire EEG assessments were performed with participants’ eyes closed. Two 5-minute periods of rest were recorded at the beginning and after the end of the 20 minutes of LMV to assess the resting state (REST PRE and REST POST). An accelerometer, positioned on the vibrator module and connected to the EEG system, allowed detecting when vibrations were on/off for synchronization with the EEG recording.

Literature indicates that LMV applied on a resting limb enhances cortical activity, with Mu rhythm ERSP at C3 serving as a marker for this activation^27,28^. Movement illusions induced by LMV amplify this cortical activity^24,27^. As our LMV conditions were optimized to induce these illusions, Mu desynchronization during the 20-minute LMV, in comparison to the resting period, was used in all participants to quantify brain activity under illusions.

### Neurophysiological and strength measurements

#### EMG recordings

After shaving and dry-cleaning the skin with alcohol, silver chloride surface electrodes (Ag/AgCl) were positioned on the belly of the right *Flexor Carpi Radialis* (FCR) with an interelectrode distance of 2 cm (center to center). To ensure consistent electrode placements across visits, the distance from the medial humeral epicondyle and the midpoint of the distal wrist crease was measured. Electrodes were positioned at 35% of this distance from the proximal reference point (participants’ mean was 9.82 ± 0.83 cm), corresponding to the theoretical optimal electrode site^39^. A reference electrode was placed over the medial epicondyle of the radial styloid. To prevent noise during LMV, the ground electrode was placed on the olecranon process. The EMG signal was amplified (gain 1000) and band-pass filtered (10-500 Hz), digitized at a sampling rate of 2000 Hz, and recorded for off-line analysis (Biopac System Inc., Goleta, CA, USA).

#### Maximum isometric grip strength

A warm-up phase consisting of 2 minutes of right wrist and finger flexions starting from low to submaximal intensity preceded maximum force evaluation. Three (n=3) maximal grip voluntary contractions (T.K.K.5401 GRIP-D hand-grip dynamometer, Takei Scientific Instruments Co., Ltd, Tokyo, Japan), lasting 3 seconds with 30 seconds recovery in between, were performed in each PRE (V1-V4) and POST (V1 & V2) visits (Figure 1). The position of the participants was similar across sessions, with the elbow and forearm resting on their right thigh while squeezing the dynamometer. Verbal encouragement was provided during every contraction. The maximal grip strength for each set of contractions was determined as the highest value among the three trials, displayed on the dynamometer screen.

#### Peripheral electrical stimulation

The H-reflex and M-wave at the right FCR were elicited by stimulating the right median nerve. Rectangular 1 ms wave pulses were delivered through bipolar felt pad electrodes, which were secured around the arm using a Velcro band, and connected to a constant-current stimulator (DS7R, Digitimer, Welwyn Garden City, Hertfordshire, UK). The optimal stimulation site, determined in every visit as the location that produced the largest M-wave, was identified near the medial border of the cubital fossa, approximately 2 cm above the medial epicondyle of the humerus. In each session, the stimulation intensity was incrementally increased until the largest H-reflex and the M-wave reached a plateau. Three stimulations were conducted at 120% of the plateau intensity, and the M-wave with the greatest peak-to-peak amplitude was designated as Mmax (mean intensity across visits: 16.0 ± 6.3 mA).

For the H-reflex, the initial visit (V1_PRE_) was used to determine a reference value: 3 stimulations were performed at the intensity eliciting the largest H-reflex; the average of these 3 peak-to-peak amplitudes was used to calculate Hmax. Subsequently, 10 stimulations were delivered at the intensity evoking 80% of Hmax (H80) within the ascending part of the recruitment curve. In the following PRE and POST visits, to ensure consistent stimulus conditions for evaluating the H80 across these different time points, a stable M-wave was maintained, corresponding to the M-wave associated to H80 (M(H80))^40^; the mean intensity across visits was 6.9 ± 3.4 mA. Only ten out of nineteen participants exhibited an H-reflex in the FCR likely due to the methodological approach of measuring the reflex in a resting muscle.

#### Transcranial magnetic stimulation (TMS)

TMS on the left primary motor cortex was delivered using a 70 mm figure-of-eight coil connected to a monophasic Magstim BiStim^2^ stimulator (The Magstim Co., Whitland, UK). The hot spot, denoted as the optimal stimulation site producing the largest Motor Evoked Potential (MEP) amplitude for a given intensity, was determined with the assistance of a navigated brain stimulation system (Brainsight TMS Navigation, Brainbox, Cardiff, UK). The hot spot, identified as one of the 12 points on the system map (MNI coordinates: -44.7, 13.8, 85.7; Négyesi et al., 2020), remained consistent across sessions. The resting motor threshold (RMT) was defined as the minimum stimulus intensity required to evoke at least 5/10 MEPs with a peak-to-peak amplitude of 50 µV in the relaxed FCR^42^. A block of 15 single-pulse was delivered at 130% of RMT to measure MEP. Another block of 15 paired-pulse stimulations at 80%-130% of RMT with 3-ms intervals was carried out to study Short Intracortical Inhibition (SICI)^43^. Three of the nineteen participants were excluded from the TMS-related measures due to either a lack of short intracortical inhibition or an excessively high resting motor threshold.

### Data analysis

#### Processing

A tailored MATLAB algorithm (MATLAB R2024a; MathWorks, Natick, MA) was developed to extract data from each stimulation (V1-V4), encompassing peak-to-peak amplitudes of Hmax, M(Hmax), H80, M(H80), Mmax, MEP, and SICI. Normalization (% expression) of Hmax, M(Hmax), H(80), M(H80), and MEP was performed relative to Mmax, facilitating intra-sessions and interindividual comparisons^40,44^.

SICI was computed with the following formulae^44^:

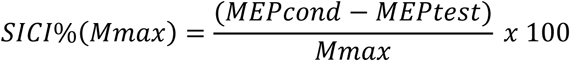

*MEP_cond_ is a MEP evoked at 130% of the RMT preceded 3 ms before by a stimulation at 80% RMT. MEPtest is a MEP evoked at 130% of the RMT*.

Negative values signify inhibition, while positive values indicate facilitation.

The interquartile range (IQR) method was employed to detect outlier values within each participant, stimulation block, and session (see supplemental Table A.1). An outlier was identified if the value exceeded Q3 + 1.5 x IQR or was less than Q1 - 1.5 x IQR^7,45^. Due to the limited dataset (i.e., only 3 values), this method was not applied to Mmax, Hmax, and M(Hmax).

During LMV, some participants exhibited a small tonic vibratory reflex (TVR)^3,46^. Complementary EMG analysis, similar to the methodology used by Amiez et al (2024a)^18^ was carried out. The results revealed an average TVR of 0.08 ± 0.05 %Mmax, ranging from 0.02% to 0.17% of Mmax (averaged of V2 and V3). These findings align with those reported in the illusions group of Amiez et al (2024a)^18^ (0.05 ± 0.03%Mmax) compared to 0.22 ± 0.18%Mmax in their TVR group. Additionally, our EMG analysis during maximal grip strength closely matches their results, as our participants demonstrated an average of 6.01 ± 3.97 %Mmax, comparable to the ∼ 6% reported their study.

#### EEG

EEG processing and calculations were performed using the MNE Python library^47^. EEG data were filtered with a 0.5 to 40 Hz band-pass filter. To improve Independent Component Analysis (ICA) decomposition, the initial 180 seconds of acquisition were excluded to eliminate noise caused by eye movements or other disruptions typically occurring during the transition to a resting state. ICA decomposition was performed and components related to eye movements and muscle artifacts were manually removed.

ERSP Analysis Event-Related Spectral Perturbations (ERSPs) were calculated using Morlet wavelets with a cycle length progressively increasing from 1 to 30, over a frequency range of 1 to 30 Hz across the full experiment duration^47^. An 80-second baseline during the resting state (-110 to -30 seconds) was applied. For analysis, only the mu band (8–13 Hz) was extracted from the ERSP data.

### Statistical analysis

#### Sample size

A power analysis was conducted using G*Power (version 3.1.9.4) to determine the required sample size for this protocol. The calculation was informed by previous studies from Souron et al. (2018, 2017c), which examined strength differences following LMV training. With a set alpha level of 5% and a statistical power of 90%, the analysis indicated that 19 participants were needed.

Statistical analyses were conducted using JASP (version 0.19.0.0 ; University of Amsterdam, The Netherlands), and “rmcorr R package” for repeated-measures correlations^48^.

Data normality was verified using the Shapiro-Wilk test.

#### Validation of the SICI protocol

One-sample t-tests (comparison with the zero ‘0’ value) were performed to confirm that the protocol successfully induced SICI during the control conditions (V1_PRE_, V1_POST_, and V2_PRE_), as described and illustrated in the supplemental Figure A.1.

#### Effects of LMV on illusions

To verify that our LMV protocol successfully induced sensory illusions, we calculated the average pooled score from the three visual analog scales over the 20-minute session and compared this score with the zero ‘0’ value (one-sample t-tests), for the second LMV session (EEG1).

We computed an ERSP_REST_ parameter that corresponds to the ERSP_MU_ normalized by the baseline ERSP_MU_ (during REST PRE) for each participant.

Then, to evaluate the acute effect of LMV on illusion, we tested the within-session evolution of sensory illusion during LMV (i.e., during one session, EEG1), as measured subjectively (pooled score) and objectively (ERSP_MU_). Two Friedman tests were performed with a factor *Time*: 1’, 5’, 10’, 15’, 19’, for pooled scores; and REST PRE, 1’, 5’, 10’, 15’, 19’, REST POST, for ERSP_MU_. The relationship between subjective (pooled scores) and objective illusion (ERSP_REST_) measures during EEG2 was further analyzed through a Pearson’s correlation for inter-participants and repeated measures correlation for intra-participant consistency^48^.

Lastly, to examine the progression of illusions across multiple LMV sessions, both intra- and inter-session changes were analyzed. For subjective pooled scores, a repeated measure ANOVA was used with a *Time* effect (1’, 5’, 10’, 15’, 19’), and a Friedman test for the *Session* effect (Sessions 1, 2, 3, 4, 5, 6, 7, 8, 9). For the ERSP_MU_ variable, a Friedman test was used with *Time* (1’, 5’, 10’, 15’, 19’) as a factor. The session effect (EEG1, EEG2) was further analyzed using a Wilcoxon test. Finally, the relationship between subjective (pooled scores) and objective illusion (ERSPREST) measures during the last LMV session (EEG2) was analyzed through a Pearson’s correlation for inter-participants and repeated measures correlation for intra-participant consistency.

#### Effects on Grip strength and neurophysiological parameters

The acute effects of LMV were assessed using two-factor repeated measures ANOVA for normally distributed variables (RMT and M(H80)), with *Visits* (V1, V2) and *PrePost* (PRE, POST) as factors. For non-normally distributed data (SICI, MEP, Mmax, H80, and Grip strength), a Friedman test was performed with a single factor, *Visits* (V1_PRE_, V1_POST_, V2_PRE_, and V2_POST_). This test was further completed by a Wilcoxon test to compare the relative changes between PRE and POST in V1 and V2, calculated as follows: V1_RATIO_ = (V1_POST_ – V1_PRE_) / V1_PRE_, and V2_RATIO_ = (V2_POST_ – V2_PRE_) / V2_PRE_.

To evaluate the chronic effects of LMV, a repeated measures ANOVA was used for normally distributed variables (RMT, M(H80), and Grip strength) across the factor *Visits* (V2_PRE_, V3, V4). For non-normally distributed data (SICI, MEP, Mmax, and H80), a Friedman test was applied with the same factor.

#### Effect size, post-hoc analyses, and significance threshold

For ANOVA tests, violations of the sphericity assumption were corrected using the Greenhouse-Geisser adjustment when necessary.

Effect sizes were calculated as follows: For ANOVA, partial Eta squared (ղ ^2^) was reported and interpreted as small (< 0.01), medium (< 0.06), and large (≥ 0.14). For the Friedman test, effect size was determined using Kendall’s W (W), categorized as small (< 0.3), medium (< 0.5), and large (≥ 0.5). For the Wilcoxon test, the rank-biserial correlation (r_B_) was computed, following the same interpretation as Kendall’s W.

Post hoc analyses were applied to all significant results: The Holm-Bonferroni was used for ANOVA, and Conover’s post-hoc test was applied for the Friedman test. Statistical significance was set at *p < 0.05* for all analyses.

## Results

### Acute impact of LMV (intra-visit effects)

#### Subjective scales of sensory illusions

Table 1 shows the average values (± SD) of the 3 visual analog scales used by participants (n=19) to subjectively rate their sensory illusions, which were recorded at specific time points through the 20-minute LMV (i.e., 1, 5, 10, 15, and 19 minutes) in the session EEG1 (i.e., the day of the first EEG recording). The majority of participants developed sensory illusions, except 2 participants. The average scores during the 20 minutes were: 3.7 ± 2.2 for strength, 5.1 ± 2.9 for continuity, and 3.9 ± 2.5 for vividness. One-sample t-test (comparison with the zero ‘0’ value) on pooled sensory illusion values showed significant differences (t = 8.25, p < 0.001). The Friedman test did not show a significant *Time* effect (χ^2^ = 2.142, p = 0.710, W = 0.028) for the evolution of the Pooled illusions overtime during the session EEG1.

**Table 1:**
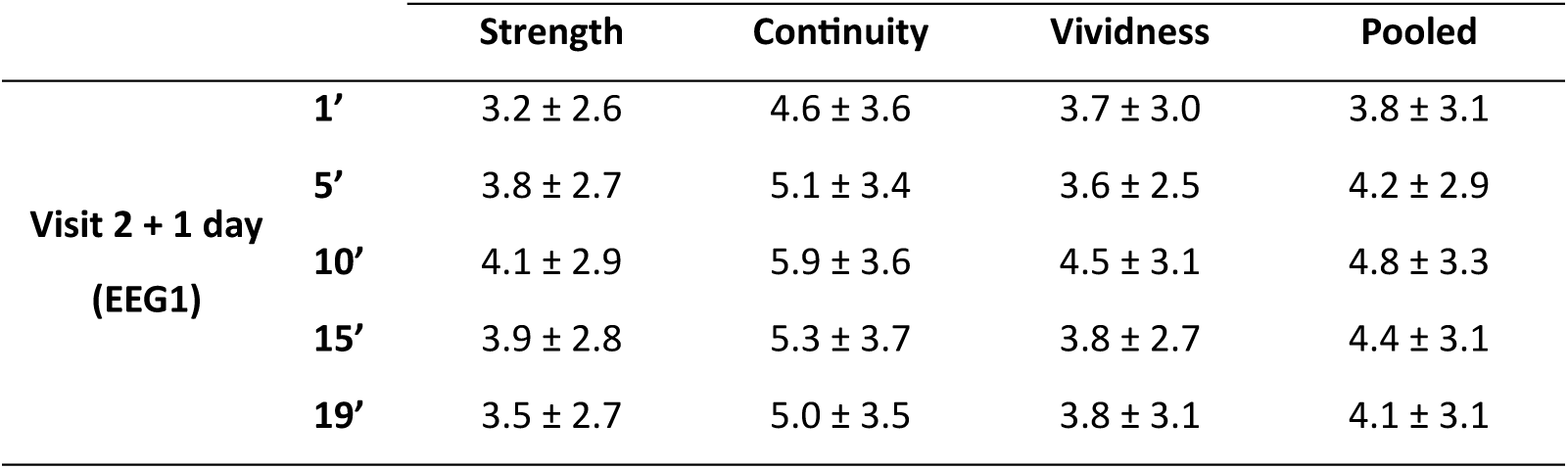
Evolution of the 3 visual analog scales used to subjectively rate the illusions perceived by participants (n=19), at specific time points during the 20-minute LMV (i.e., 1, 5, 10, 15, 19 minutes), during EEG1 (on the same day as the first EEG recording).

#### EEG analysis during LMV

The EEG analysis at the EEG1 session indicated a large desynchronization of the ERSP_MU_ over the 20-minute LMV, supported by a significant *Time* effect (χ^2^ = 51.338, p < 0.001, W = 0.450; see Figure 3). Post-hoc analysis did not show a significant difference between REST PRE and REST POST (p = 0.56), or REST PRE and 19’ (p = 0.17). Significant modifications, however, were found between REST PRE and 1’ (p < 0.001), 5’ (p < 0.001), 10’ (p < 0.001), and 15’ (p = 0.003), and between 1’ and 15’, 19’, and REST POST (for all tests, p < 0.001).

**Figure 3:**
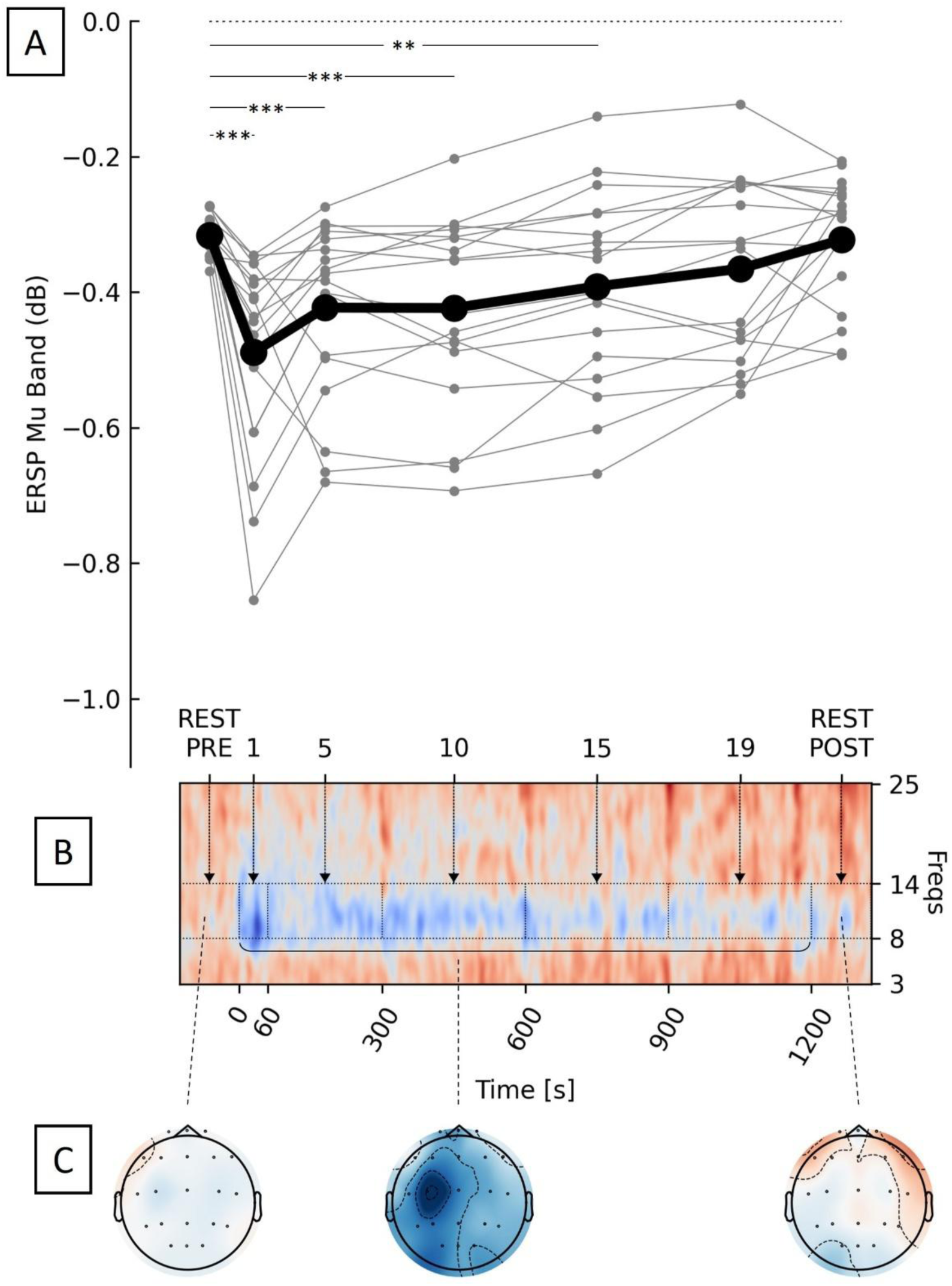
Illustrations of the alpha band evolution before (REST PRE, from -120s to 0s), during, and after LMV (REST POST from 1201s to 1440s) in EEG1. Figure 3A shows the mean of ERSPMU at different time points: REST PRE, 0-60s (1), 61-300s (5), 301-600 (10), 601-900s (15), 901-1200s (19), and REST POST. Individual data are represented in grey and the mean of all participants (n=19) in black. p: *** ≤ 0.001, ** ≤ 0.01. Figure 3B represents the heatmap (mean of all participants) for frequencies from 3 to 25 Hz. The blue color illustrates a desynchronization. Figure 3C shows the scalp maps (of one representative participant) during the REST PRE (on the left), during LMV (on the middle), and the REST POST (on the right).

#### Correlation between subjective sensory estimation and *ERSP_MU_*

Inter-participant correlations were calculated between the pooled subjective scores provided by the participants during the 20-minute LMV period and the mean recorded ERSP_REST_ during the session EEG1. The correlation did not reach significance (r = 0.29, p = 0.11, supplemental Figure A.2A). A repeated measure correlation analysis did not show any significant intra-participant correlation between the evolution of ERSP_REST_ and the subjective pooled illusions scores during EEG1 (r = 0.089, p = 0.444, supplemental Figure A.2B). This indicates that the subjective scores and the desynchronization patterns during LMV are independent.

#### Neurophysiological and strength evaluations

Figure 4 shows the average values of all the parameters recorded in V1 and V2. For the H-reflex (H80, Figure 4A), statistical analysis revealed a significant *Visits* effect (χ^2^ = 8.40, p = 0.038, W = 0.280). Post-hoc comparisons indicated a significant reduction in H-reflex amplitude (p = 0.028) between V1_POST_ (after a 20-minute rest) and V2_POST_ (after a 20-minute LMV), while no other comparisons reached significance (p ≥ 0.133). The Wilcoxon test, comparing V1_RATIO_ (mean across participants: -6.2 ± 30.4%) and V2_RATIO_ (-45.3 ± 37.9%) revealed a significantly larger reduction of the H80 during V2 compared to V1 (p = 0.020, r_B_ = 0.818). A qualitative illustration of H-reflex reduction is depicted in Figure 5A, which displays representative H-reflex waveforms from a typical participant in both V1 and V2.

**Figure 4:**
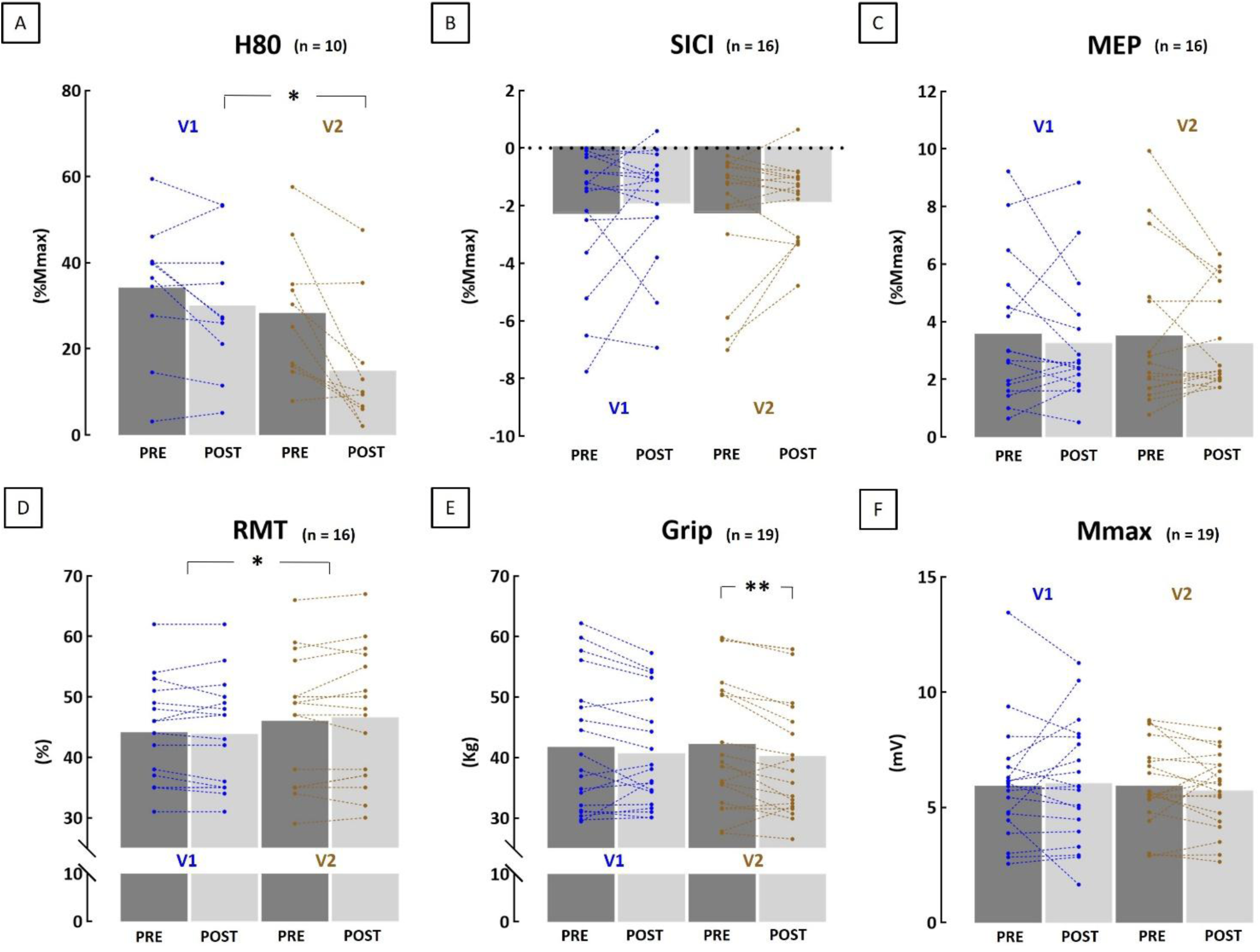
PRE and POST values in V1 (after 20-minute rest) and V2 (after 20-minute LMV). The mean values of all participants are represented in dark grey columns for the PRE values and in light grey columns for the POST values. The blue and brown lines and plots represent the evolution of the individual data between PRE and POST, V1 and V2 respectively. Statistical differences are indicated with p: ** < 0.01 and * < 0.05. H80 = 80% of maximal H-reflex, SICI = short intracortical inhibition, MEP = Motor evoked potential, RMT = resting motor threshold, Mmax = maximal M-wave, %Mmax = expressed as a percentage of maximal M-wave.

**Figure 5:**
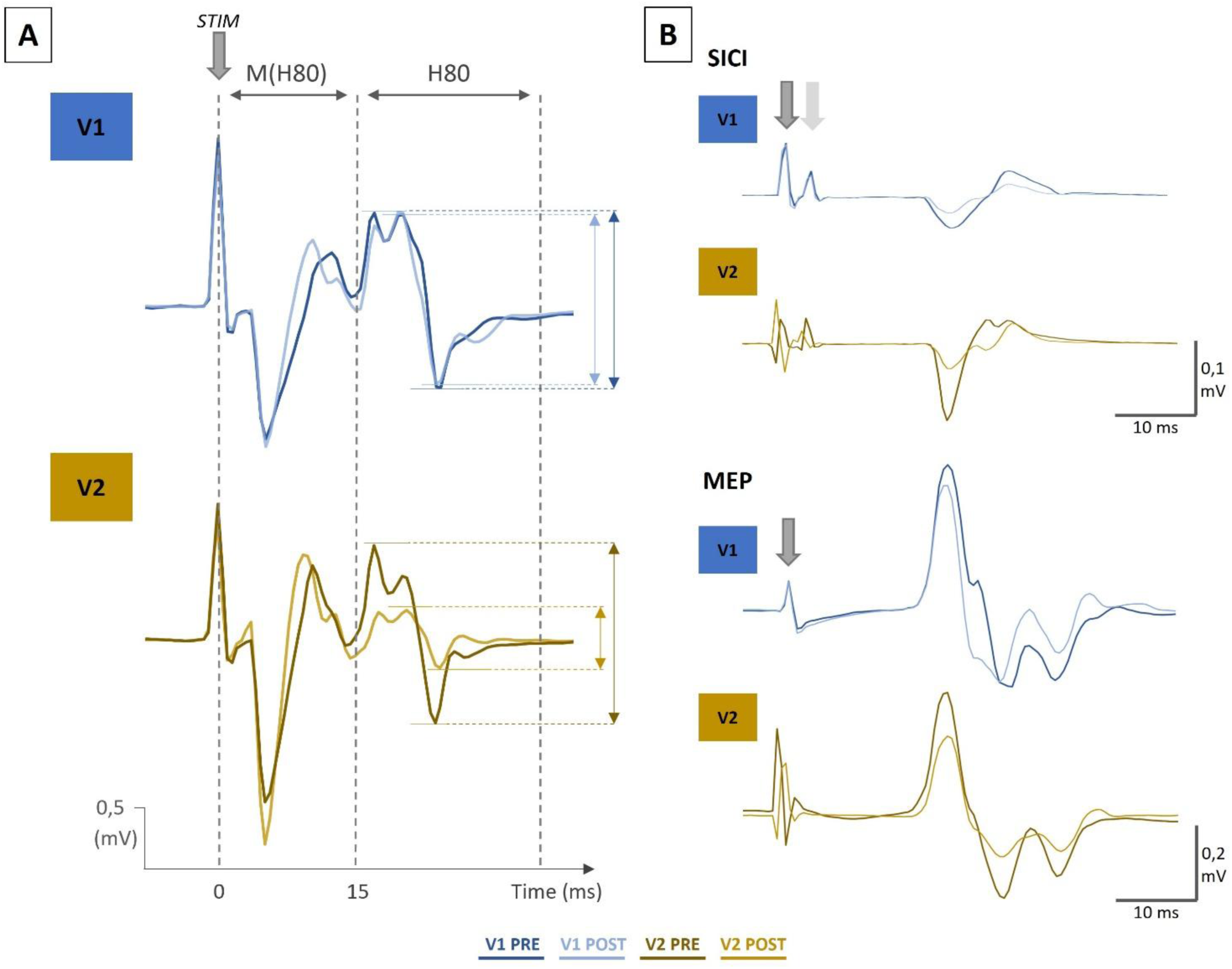
Traces for the H80 (Fig. 5A), SICI, and MEP (Fig. 5B) parameters for the PRE and POST sessions in V1 (after 20 minutes rest) and V2 (after 20-minute LMV) from a typical participant. In blue for V1PRE and V1POST, and brown for V2PRE and V2POST.

The neurophysiological parameters presented in Figures 4B, 4C, and 4F showed no significant effects, including SICI (χ^2^ = 0.375, p = 0.945, W = 0.008, see Figure 5B), MEP (χ^2^ = 0.975, p = 0.807, W = 0.020 see Figure 5B), and Mmax (χ^2^ = 0.537, p = 0.911, W = 0.009), and no V1_RATIO_ and V2_RATIO_ differences (p ≥ 595, r_B_ ≤ 0.162). For RMT (Figure 4D), no *Visits * PrePost* interaction or *PrePost* effect was detected (F ≤ 1.215, p ≥ 0.288, ղ ^2^ ≤ 0.075). However, a significant main effect of *Visits* was identified (F = 6.680, p = 0.021, ղ ^2^ = 0.308), indicating a slight increase in RMT in V2 in comparison to V1.

Regarding grip strength, a significant effect was found (χ^2^ = 8.952, p = 0.030, W = 0.157). Post-hoc analyses revealed a significant reduction in strength between V2_PRE_ and V2_POST_ (p = 0.007), while no other comparisons reached significance (p ≥ 0.127). However, the Wilcoxon test comparing V1_RATIO_ (mean across participants: -1.2 ± 8.6%) and V2_RATIO_ (-4.3 ± 7.5%) showed no significant difference in grip strength during V2 compared to V1 (p = 0.459, r_B_ = 0.205).

A detailed statistical analysis is presented in the supplemental Table A.2.

The acute effects observed in V2 following LMV, compared to the V1 control condition, align with previous findings in the literature. Specifically, we found a reduction in H-reflex amplitude^4,7^, no significant changes in MEP, SICI, or Mmax^7,17^, and a slight decrease in grip strength, though, not exceeding the reduction observed in the control condition^18^. These results confirm that our LMV application meets the standard conditions.

### Chronic impact of LMV (inter-visit effects)

#### Subjective scales of sensory illusions

Figure 6 shows, for the 9 vibratory sessions (V2 to EEG2), the average pooled score of the 3 visual analog scales used to subjectively rate the sensory illusions perceived by participants (n=19). The majority of participants felt sensory illusions (n=17), except for 2 participants throughout the entire protocol. On average, sensations were rated at 3.8 ± 2.5 for strength, 5.1 ± 3.1 for continuity, and 3.9 ± 2.6 for vividness over the 20 minutes across all LMV sessions. The repeated measure ANOVA revealed a significant *Time* effect (F = 34.819, p < 0.001, ղ ^2^ = 0.813, Figure 6). Post-hoc analyses demonstrated significant differences between 1’ and 5’ (p < 0.001), 10’ (p < 0.001), 15’ (p < 0.001) and 19’ (p = 0.011), between 10’ and 19’ (p = 0.007), and between 15’ and 19’ (p = 0.020). A Friedman test did not reveal any significant *Session* effect (χ^2^ = 7.164, p = 0.519, W = 0.047), indicating that the illusions remain consistent across sessions.

**Figure 6:**
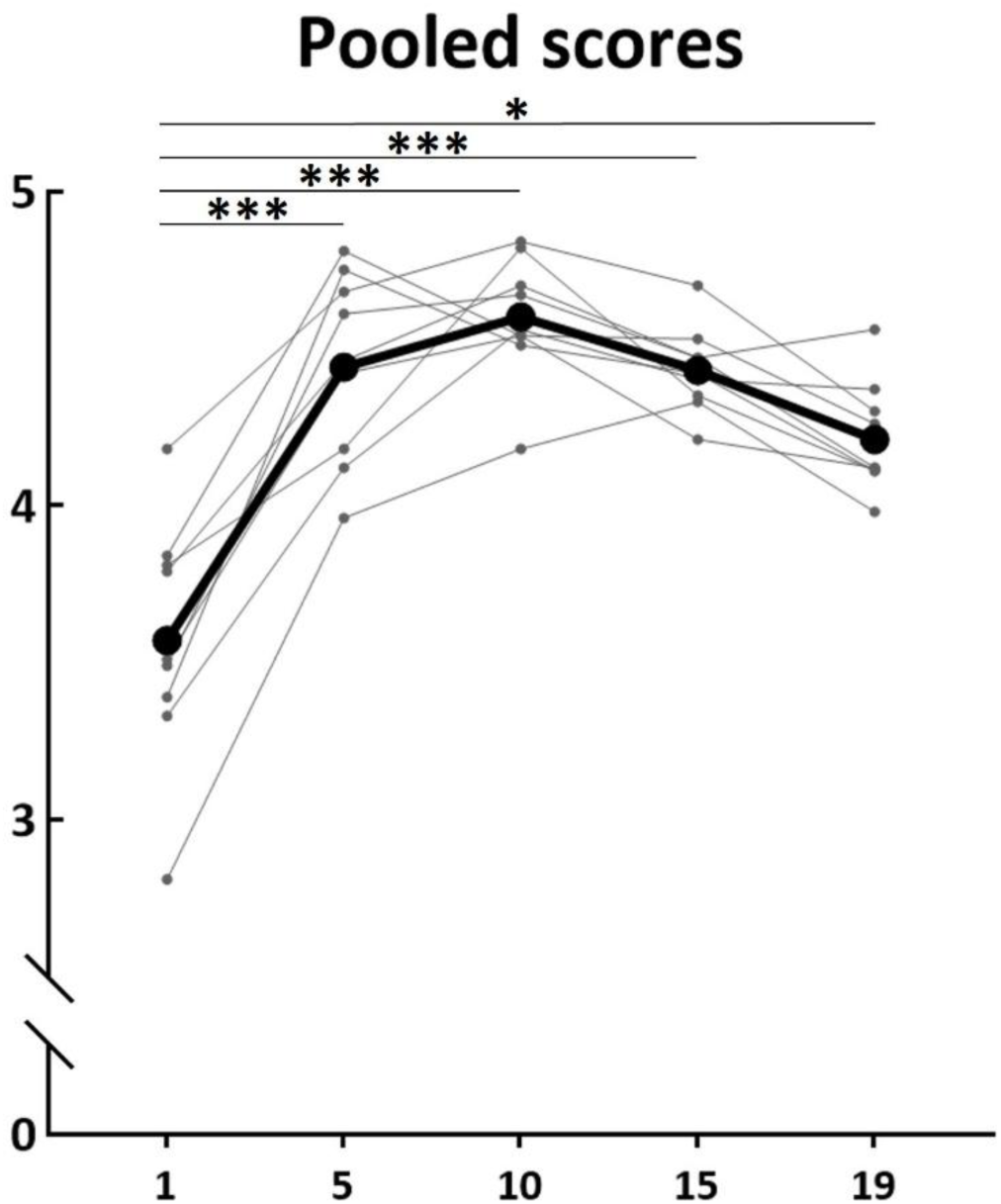
Evolution of the Pooled scores (average of the 19 participants) at specific time points (1, 5, 10, 15, and 19 minutes) over the 9 vibratory sessions. In grey, the pooled scores by session (from V2 to EEG2), and in black the average of the 9 vibratory sessions. p: *** < 0.001 and * < 0.05.

#### EEG analysis during LMV

The analysis of the EEGs recorded at the EEG1 and EEG2 sessions revealed a large desynchronization of the ERSP_MU_ over the 20-minute LMV. Similar to the EEG1 session, there was a significant *Time* main effect (χ^2^ = 16.707, p = 0.002, W = 0.220) in the EEG2 session. Post-hoc showed significant modifications between 1’ and 15’, 19’ (for all analyses, p ≤ 0.011), between 5’ and 19’ (p = 0.017), and between 10’ and 19’ (p = 0.010), confirming the strong desynchronization of the alpha band mainly during the first 10 minutes of both the first and last LMV sessions. No difference was found between the two sessions (p = 0.953, r_B_ = -0.021).

#### Correlation between subjective sensory estimation and *ERSP_MU_*

Similar to EEG1, no significant inter-participant correlations were found between the average subjective pooled scores reported during the 20-minute LMV period in EEG2 and the mean ERSP_REST_ values recorded during the same session (r = 0.004, p = 0.988). The repeated measure correlation analysis also revealed no significant intra-participant correlation between the evolution of ERSP_REST_ and subjective illusion pooled scores during EEG2 (r = -0.053, p = 0.647).

#### Neurophysiological and strength evaluations

Figure 7 shows the average and individual values of all the evaluated parameters for V2_PRE_ (the baseline), V3 (after the 9 sessions of LMV), and V4 (5 days after the end of the LMV protocol). None of the evaluated parameters displayed any chronic adaptation due to LMV. This was confirmed by the absence of *Visits* main effect (for all analyses, F ≤ 1.978, p ≥ 0.156, ղ_p_^2^ ≤ 0.153; χ^2^ ≤ 3.125, p ≥ 0.210, W ≤ 0.098). Statistical details are presented in supplemental Table A.4, and the evolution of M(H80) is presented in supplemental Figure A.3.

**Figure 7:**
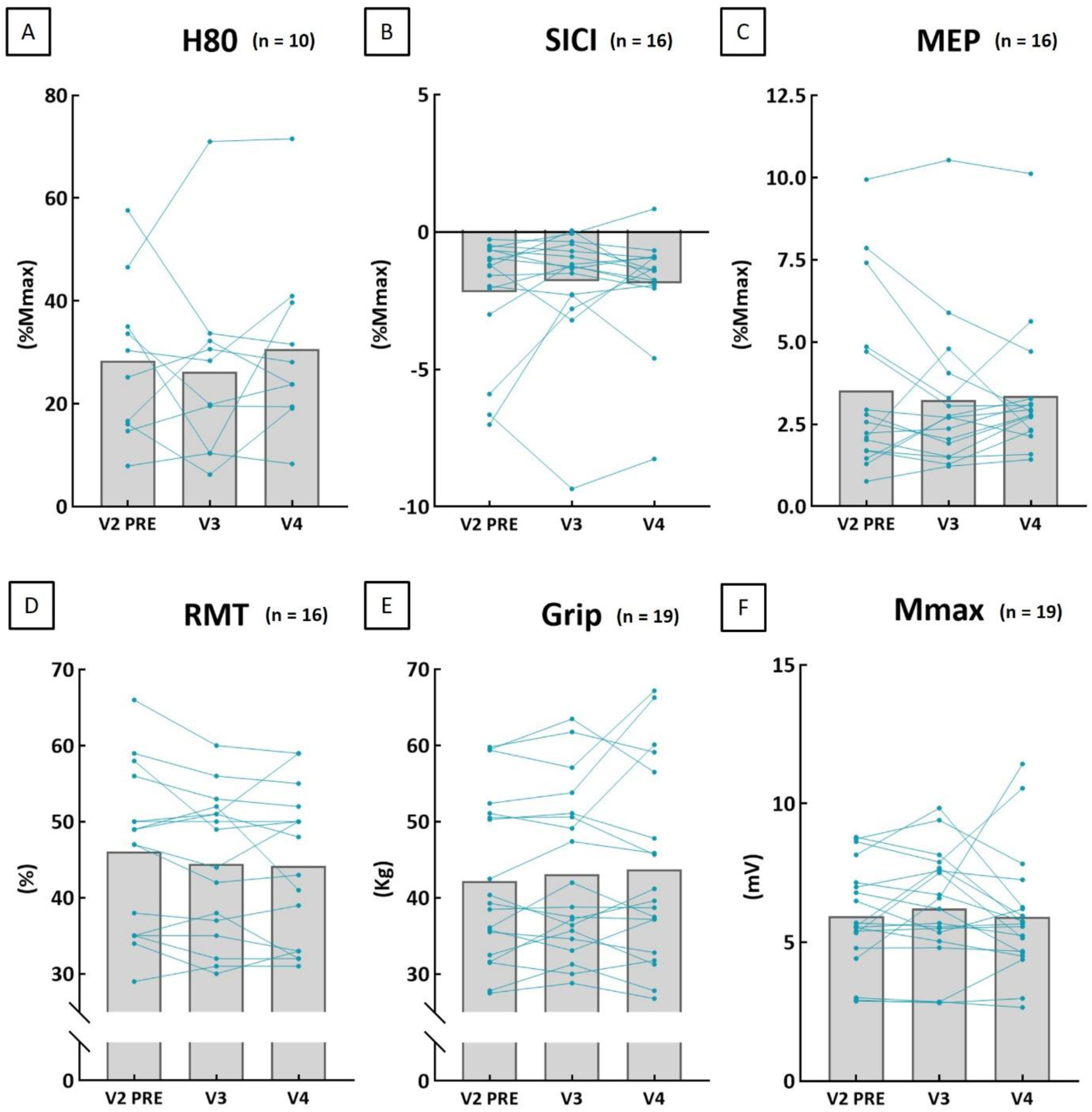
Evolution of the neurophysiological parameters for V2PRE (the baseline), V3 (following the 9 LMV sessions), and V4 (5 days after the end of the LMV). Individual data are represented in blue and the mean of all participants is represented by the grey column. H80 = 80% of maximal H-reflex, SICI = short intracortical inhibition, MEP = Motor evoked potential, RMT = resting motor threshold, Mmax = maximal M-wave, %Mmax = expressed as a percentage of maximal M-wave.

## Discussion

In this study, we investigated the neurophysiological and functional effects of nine sessions of LMV training designed to induce movement illusions. We assessed spinal, corticospinal, and intra- cortical excitabilities, as well as grip strength, of the vibrated FCR muscle in 19 healthy participants. Our results attest that the LMV protocol induced movement illusions, as quantified by answers to a subjective questionnaire and by increased cortical activity (i.e., desynchronization of the Mu-band, ERSP_MU_ measured by EEG). At the acute level, our results aligned with the existing literature; that is a single LMV session significantly reduced spinal excitability^4,7^ without significantly changing muscle strength^18^, short intracortical inhibition, and cortical excitability (MEP)^17^, when compared to the control condition. Following the 9 sessions of LMV conducted across 11 days, no chronic modulations in intracortical or corticospinal excitabilities were observed, nor were there significant changes in grip strength. To our knowledge, this is the first study to examine the long-term effects of LMV on the upper limb under controlled conditions aiming to elicit movement illusions.

In our study, both subjective (i.e., using visual analog scales) and objective (i.e., EEG recording) measures were employed to evaluate movement illusions. Most participants (17 out of 19) reported experiencing illusions across the 9 LMV sessions, and all participants exhibited Mu band desynchronization over the sensorimotor cortex (C3), consistent with previous studies^14,27,28^. Our findings indicate that illusions can persist over extended durations, with the strongest sensations reported between 5 and 15 minutes of LMV. Correspondingly, cortical activity was significantly heightened during the first 15 minutes of LMV. This aligns with a previous study that assessed wrist illusions using the same subjective scales over 30 minutes of continuous LMV, observing sustained illusions throughout the session with peak scores at the beginning, and lessening toward the end^7^.

To our knowledge, this study is the first to repeatedly assess movement illusions using both methods. Interestingly, no significant correlation was found between these two variables, suggesting they may reflect distinct underlying processes. While we could assume that subjective illusions are directly linked to cortical activity during LMV, our findings challenge this notion. One possible explanation is that the absence of a vibration condition without illusions limited our ability to establish a clearer relationship between these measures. Nonetheless, the significant effect of time observed for both variables indicates that meaningful fluctuations were captured, reinforcing the idea that they may represent independent processes.

Note that complementary correlation analyses were conducted to explore the relationship between movement illusions and the different neurophysiological and strength measures, both acutely (see supplemental Table A.3) and chronically (see supplemental Table A.5). A strong inverse correlation was identified between subjective illusion ratings (across the mean scores of all scales: pooled scores) and the absence of a tonic vibratory reflex (TVR, see supplemental Table A.3), consistent with the existing literature^3,18,20^. However, no such association was observed between ERPS_REST_ desynchronization and the absence of a TVR. It is worth noticing that the EEG evaluation was conducted 1 day (EEG1) after the TVR measurement (V2), which may have influenced the observed results.

As previously mentioned, two participants did not experience any movement illusions throughout the experiment. To ensure the robustness of our findings, all correlations and analyses presented in the paper were re-evaluated after excluding these two subjects. The outcomes of the study remained unchanged, with the exception of the correlation between TVR and illusions (pooled score), which was no longer significant.

Our protocol successfully replicated a well-documented effect of acute LMV on spinal excitability, namely a significant reduction in the H-reflex amplitude (-45.3 ± 37.9%) immediately following LMV^4,7,10^. This reduction in spinal loop excitability is consistent across studies, irrespective of the specific LMV conditions that are employed^4^. No other significant acute effects of LMV were observed in our study. Consistent with prior research, we did not detect acute changes in RMT and Mmax, two parameters known to remain stable over time^7,19^.

Similarly, our findings align with studies employing LMV conditions similar to ours, which reported no acute effects on MEP amplitude^11,12,15^. Note however that a recent study has reported a MEP reduction following LMV with illusion^7^. It has been suggested that when measured by TMS, possible corticospinal increase may be masked by a reduction in motoneuronal excitability. Notably, some studies have reported acute increases in the MEP/CMEP^7,11^ or MEP/TMEP^12^ ratios, indicating a transient rise in cortical excitability during the first 30 minutes following acute LMV. As our protocol did not evaluate motoneuronal excitability, we cannot confirm or refute these findings.

Our evaluation of short intracortical inhibition (SICI) revealed no acute effect of LMV. This aligns with findings from analogous LMV protocols that did not specifically control for movement illusions^15,17,49^. As SICI is strongly influenced by MEP^50^, we calculated the SICI/MEP ratio to account for potential masked effects (see supplementals Table A.2). Our analysis revealed no significant modifications of this ratio, suggesting that the observed results are not confounded by variations in MEP amplitude. These observations challenge hypotheses suggesting intracortical changes involving GABAergic circuits, particularly GABA_A_ pathways, as a mechanism for vibration-induced modulation of corticospinal excitability^4,30,49^.

Finally, we observed a slight reduction in strength post-vibrations (-4.3 ± 7.5%), which was not significantly greater than the reduction observed under resting conditions (-1.2 ± 8.6%). This finding corroborates with those of some previous studies^4,51,52^ but differs from others^4,53^. Notably, two studies with a similar LMV protocol and controlling for movement illusions, reported, for the first one, a significant acute reduction in vibrated muscle’s strength (-11.8%)^7^, however, it lacked a control condition for comparison. The second study found no significant difference in maximal strength post-vibration condition with illusions whereas a marked reduction in condition without illusions but with TVR^18^.

No chronic adaptations to LMV were observed in our study. We found that repeated exposure to LMV induced no neurophysiological adaptations at the spinal level (i.e., H-reflex) in the vibrated muscle. This finding adds to the limited literature, where studies have reported increased^31,32^ or unchanged^34^ H-reflex amplitudes following LMV training. These discrepancies may arise from variations in LMV parameters (i.e., 50 Hz for 2 weeks, 100 Hz for 8 weeks, and 80 Hz for 2 weeks, in the studies by Lapole et al (2013)^31^, Souron et al (2017)^34^ and ours, respectively), evaluation methodologies (Lapole (2013)^31^ did not adjust with M-wave (associated to the H-reflex) equalization, Souron (2017)^34^ evaluated H-reflex during muscle contraction), duration application (1 hour per day for Lapole and Souron studies, and 20 min in ours), or the targeted segment (i.e., Achilles’ tendon for Lapole, tibialis anterior for Souron, wrist in our study).

As expected, maximal M-wave and RMT remained stable across visits, consistent with previous findings indicating these parameters are unaffected by time or LMV^19,32,34^. Moreover, we did not reveal any training effects on corticospinal excitability (MEP), aligning with prior works reporting no significant changes following LMV training^33,34^. However, the possibility of cortical adaptations cannot be entirely dismissed. A more comprehensive approach incorporating an evaluation of motoneuronal excitability, as described in the previous paragraph, could provide deeper insights.

Furthermore, no chronic impact of LMV on SICI was observed. This is the first study that measures the chronic effect of LMV on intracortical inhibition, as measured by SICI. Our results align with those of a previous study reporting no effect on the silent period duration^34^, but are in contrast with another work that found a reduction in this silent period^33^. The interpretation of these results is complicated by the fact that the methodology used to assess intracortical inhibition is influenced by confounding mechanisms that make it challenging to isolate and analyze^54^. To rule out masked effects, as performed in the previous acute part, we verified the SICI/MEP ratio, but no significant changes were detected (see supplemental Table A.4). Despite increased cortical activity in sensorimotor areas during LMV, our results did not reveal any lasting effects on cortical or intracortical mechanisms.

Lastly, our results showed no significant strength increase over time, diverging from prior training studies that reported strength gains^31–34^. Although not significant, our findings did indicate a slight tendency toward increased strength (V2_PRE_ to V3: 2.7 ± 8.0%, and V2 _PRE_ to V4: 3.3 ± 10.5%). This difference may be partly attributed to the shorter duration of our protocol (under 2 weeks), which may not suffice to induce neural adaptations and strength gains^4,55^, as demonstrated in studies with longer training durations. Also, our protocol did not elicit any acute effects of LMV on strength, suggesting that LMV applied under movement illusion conditions may influence strength differently than other LMV protocols, potentially leading to distinct training adaptations^18^. These findings highlight the need for further research to explore the chronic effects of LMV under controlled conditions, particularly those designed to specifically induce tonic vibratory reflex (TVR).

It is essential to consider that this type of strong sensory stimulation alone may not be sufficient to induce long-term plasticity without the integration of cognitive or motor tasks, as suggested by some studies using congruent action observation^30,56–58^. This warrants the need for further research to explore how cognitive engagement or task-specific activities might enhance the effectiveness of LMV protocols aimed at promoting neuroplasticity.

LMV are widely used in clinical settings with numerous studies reporting their acute effects in chronic patients, such as a temporary reduction of spasticity. Several reviews provide valuable insights into their therapeutic applications^5,6,59–62^. However, the substantial variability in protocol parameters often leads to differing neurophysiological and clinical outcomes^62^. Accordingly, the underlying mechanisms of these effects remain insufficiently understood.

It is important to consider that studies conducted on healthy individuals using this type of training protocol may fail to induce chronic adaptations, as these participants already operate at their maximal capacities. Despite the relatively high LMV dose applied in our protocol (i.e., 20 minutes per day), this alone may not be sufficient to induce measurable neurophysiological or strength modulations. However, in pathological populations, where sensorimotor impairments are prevalent, such interventions may hold significant therapeutic potential to restore function and compensate for neuromuscular deficits.

Study limitations have to be noted. We selected the flexor carpi radialis (FCR) muscle due to its accessibility and the relevance of assessing upper limb extremity; however, this small muscle is known to produce relatively small MEP/Mmax ratios, often below 10% at rest^7^. This characteristic may contribute to higher variability in SICI measurements^44^. Indeed, our neurophysiological results exhibited variability, similar to other studies^17^, suggesting that a larger sample size might be necessary to strengthen the statistical power of our findings.

Only ten out of nineteen participants exhibited an H-reflex in the FCR, likely due to the methodological approach of measuring the reflex in a resting muscle—a condition that is suboptimal for eliciting this response^63^. However, this method was intentionally selected to assess excitabilities in a resting state, allowing us to investigate whether LMV could modulate the baseline neural activity of healthy participants.

Additionally, our results on grip strength should be interpreted with caution. Grip strength reflects the combined activity of multiple wrist and hand muscles, including moderate wrist extensors, not just the wrist flexors^64^. A more targeted assessment focusing exclusively on wrist flexors might have yielded more pronounced results. Furthermore, existing literature suggests that movement illusions induced by LMV may have a significant impact on the antagonist muscles involved in the perceived movement^7,26,65^. Exploring this phenomenon in future studies could provide valuable insights.

## Acknowledgments

Authors sincerely thank Corentin Delphin and Delphine Besson for their assistance in conducting the vibratory sessions, and Nicolas Frenot for his contributions to figure design. Special thanks to William Dupont for his help in training in neuronavigation and TMS, and to Yves Ballay for his support in maintaining the experimental equipment. We also extend our gratitude to Bioserenity for providing the EEG materials and, most importantly, to all the participants for their time and commitment to this study.

## Author contributions statement

The project was conceived by SJ, CP, JG and AM. SJ conducted all the experiments, SJ, JG and CP analyzed the results. All authors reviewed the manuscript.

## Data availability statement

The corresponding author can provide the datasets from this study upon reasonable request.

## SUPPLEMENTALS MATERIAL

**Table A.1:** Mean number of stimulations analyzed per variable and participant at each visit, following the removal of outlier values, calculated with the IQR method. Results are presented as mean ± SD.

| Measure | Number<br>stimulations<br>/ session | V1<sub>PRE</sub> | V1<sub>POST</sub> | V2<sub>PRE</sub> | V2<sub>POST</sub> | V3 | V4 | Total loss<br>(%) |
| --- | --- | --- | --- | --- | --- | --- | --- | --- |
| **SICI**<br>(n = 16) | 15 | 13.6 ± 1.6 | 13.8 ± 1.2 | 14.1 ± 0.9 | 13.6 ± 1.5 | 13.8 ± 1.3 | 13.7 ± 1.2 | 8.4 ± 3.4 |
| **MEP**<br>(n = 16) | 15 | 13.8 ± 1.2 | 14.0 ± 1.0 | 14.0 ± 1.4 | 13.8 ± 1.6 | 13.8 ± 1.1 | 13.8 ± 1.1 | 7.8 ± 3.0 |
| **M(H80) & H80**<br>(n = 10) | 10 | 8.2 ± 1.5 | 8.8 ± 1.6 | 9.0 ± 0.8 | 8.7 ± 1.2 | 8.1 ± 1.4 | 8.7 ± 0.8 | 14.2 ± 4.6 |

### SICI evaluation

Supplemental Figure A.1 qualitatively illustrates Motor Evoked Potentials (MEPs) obtained from single and double (SICI) TMS stimulations. One-sample t-test comparing SICI values with the ‘0’ zero value revealed significant differences for V1_PRE_ (t = -3.735, p = 0.002), V1_POST_ (t = -3.732, p = 0.002), and V2_PRE_ (t= -3.888, p = 0.001). This finding indicates that the protocol used induced intracortical inhibition.

**Supplemental Figure A.1:**
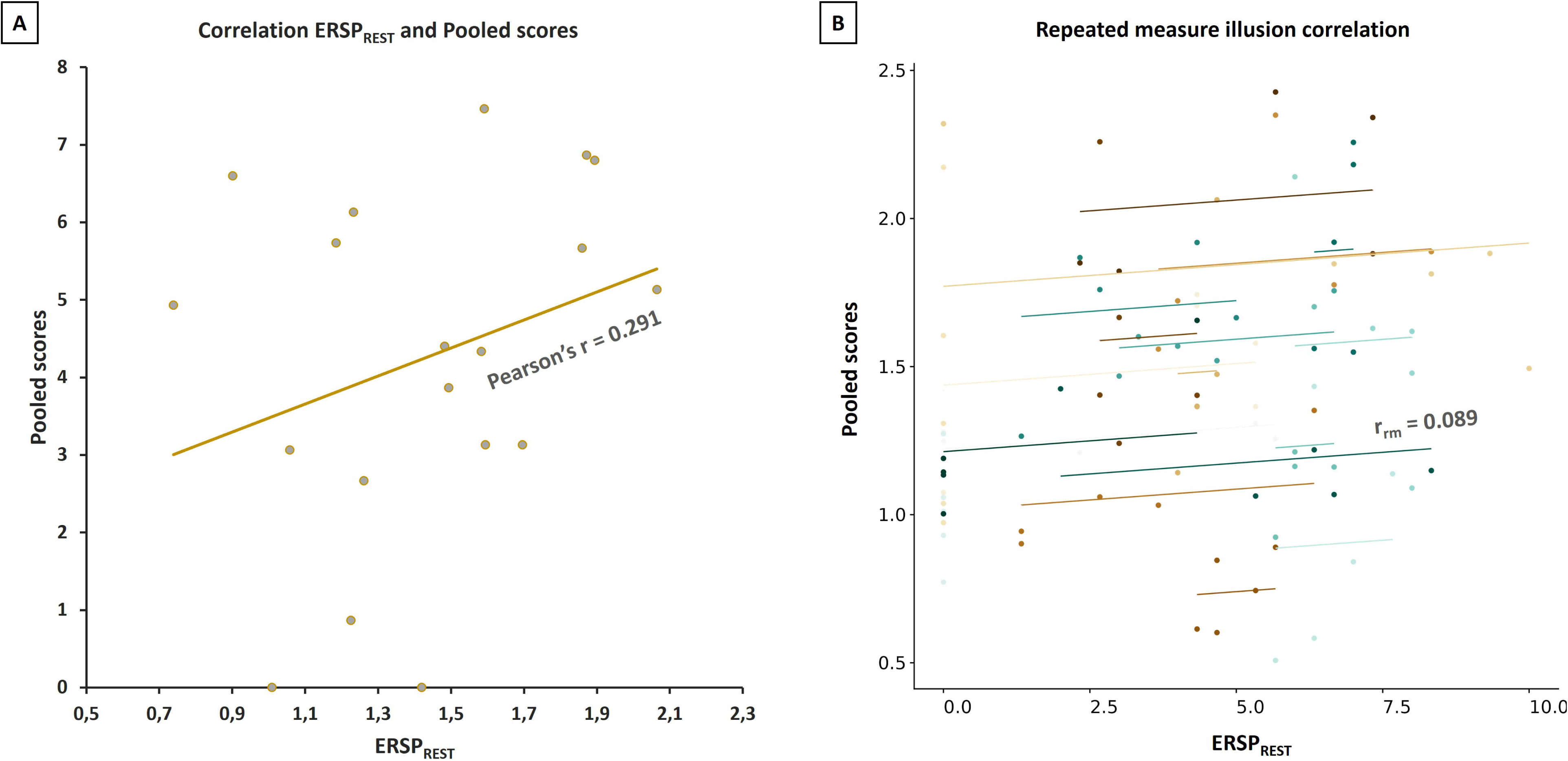
Motor evoked potential (MEP) after a single stimulation by TMS at 130% RMT (black), and after two stimulations at 80-130% RMT (SICI, in grey), are shown from one participant during V1_PRE_.

### Acute effects

#### Correlation between subjective sensory evaluation and ERSP_REST_

No correlation was observed between ERSP_REST_ and the subjective pooled scores (r = 0.29, p = 0.11). Repeated multiple correlation analysis did not show a significant intra-participant correlation between the evolution of ERSP_REST_ and the subjective illusion pooled scores during EEG1 (r = 0.089, p = 0.444).

**Supplemental Figure A.2:**
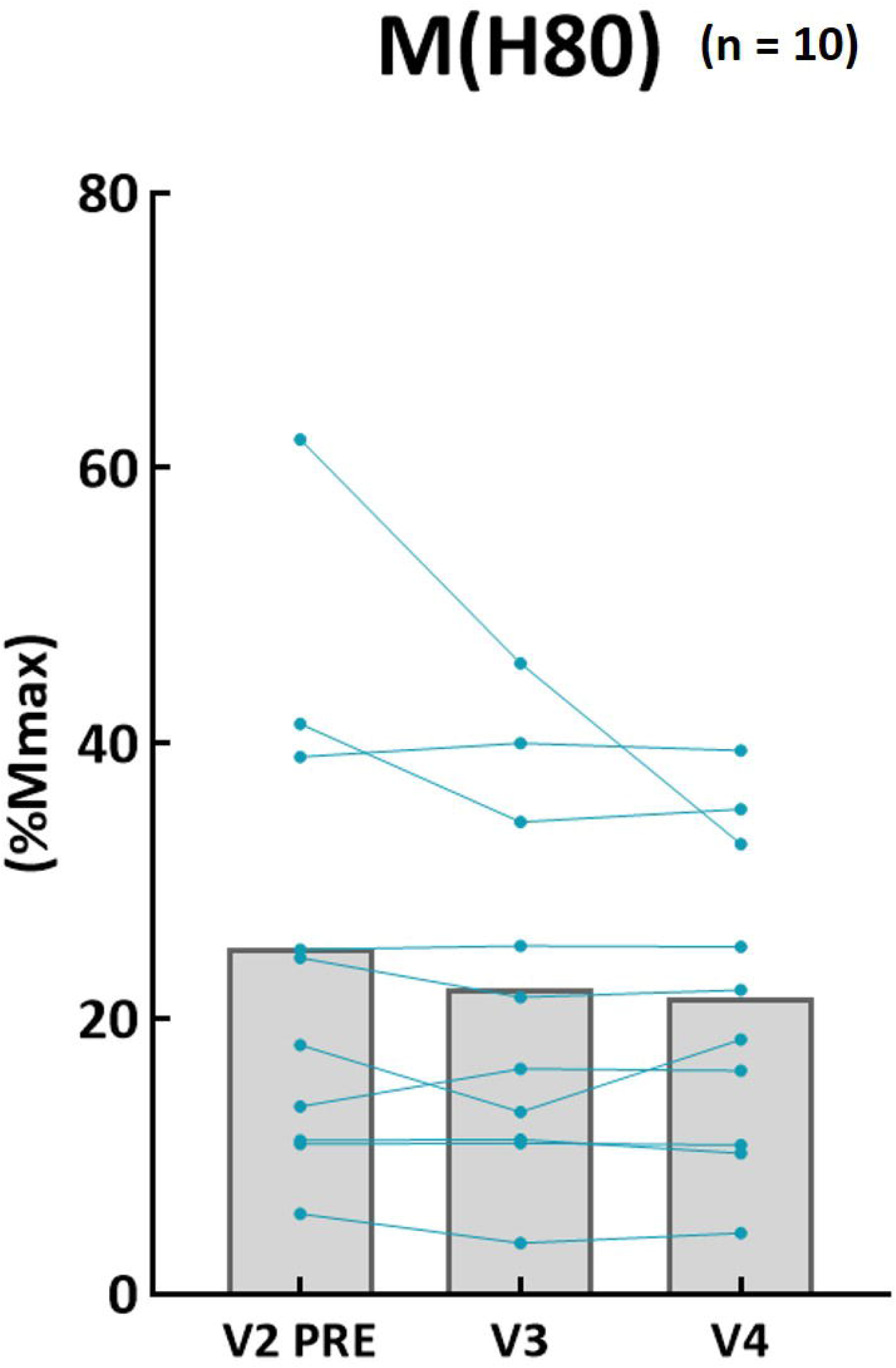
Figure A displays the inter-participant correlation between the mean subjective scores (Pooled) and the mean objective ERSP_REST_ (EEG) across all participants (n = 19) during the second LMV session (EEG1). Figure B illustrates the intra-participant repeated measure correlation between the subjective pooled scores and ERSP_REST_ for all participants (n = 19) across the entire 20-minute LMV of EEG1, at different time points (1’, 5’, 10’, 15’, and 19’).

**Supplemental Table A.2:** Results for the acute impact of LMV. Two-factor repeated measures ANOVA, with Visits (V1, V2) and PrePost (PRE, POST) as factors, or Friedman test with a single factor, Visits (V1_PRE_, V1_POST_, V2_PRE_, V2_POST_), completed by a Wilcoxon test comparing V1_RATIO_ and V2_RATIO_. p ≤ 0.05 is significant.

|  | *Visits* |  |  | *Post-Hoc* | *Wilcoxon* |  |
| --- | --- | --- | --- | --- | --- | --- |
| | $\chi^2$ | *p* | *W* | | *p* | *r<sub>B</sub>* |
| **SICI** | 0.375 | 0.945 | 0.008 |  | 0.940 | -0.029 |
| **MEP** | 0.975 | 0.807 | 0.020 |  | 0.597 | -0.162 |
| **SICI/MEP** | 0.375 | 0.945 | 0.008 |  | 0.980 | 0.015 |
| **H80** | 8.400 | **0.038** | 0.280 | V1<sub>PRE</sub>-V1<sub>POST</sub> : 0,445<br>V2<sub>PRE</sub>-V2<sub>POST</sub> : 0,133<br>V1<sub>PRE</sub>-V2<sub>PRE</sub> : 0,133<br>**V1<sub>POST</sub>-V2<sub>POST</sub> : 0,028** | **0.020** | 0.818 |
| **Mmax** | 0.537 | 0.911 | 0.009 |  | 0.595 | 0.147 |
| **Grip** | 8.952 | **0.030** | 0.157 | V1<sub>PRE</sub>-V1<sub>POST</sub> : 0,286<br>**V2<sub>PRE</sub>-V2<sub>POST</sub> : 0,007**<br>V1<sub>PRE</sub>-V2<sub>PRE</sub> : 0,127<br>V1<sub>POST</sub>-V2<sub>POST</sub> : 0,841 | 0.459 | 0.205 |

|  | *Visits* |  |  | *PrePost* |  |  | *Interaction* |  |  | *Post-Hoc* |
| --- | --- | --- | --- | --- | --- | --- | --- | --- | --- | --- |
| | *F* | *p* | $\eta_p^2$ | *F* | *P* | $\eta_p^2$ | *F* | *p* | $\eta_p^2$ | |
| **RMT** | 6.680 | **0.021** | 0.308 | 0.394 | 0.539 | 0.026 | 1.215 | 0.288 | 0.075 | **V1-V2 : 0.021** |
| **M(H80)** | 2.271 | 0.166 | 0.201 | 0.003 | 0.958 | <0.001 | 4.844 | 0.055 | 0.350 | V1<sub>PRE</sub>-V1<sub>POST</sub> : 0.490<br>V2<sub>PRE</sub>-V2<sub>POST</sub> : 1.000<br>V1<sub>PRE</sub>-V2<sub>PRE</sub> : 0.430<br>V1<sub>POST</sub>-V2<sub>POST</sub> : 1.000 |

**Supplemental Table A.3:** Results of the linear Pearson’s correlation between the mean subjective pooled scores recorded during V2, or the objective measure (mean ERSP_REST_) obtained during EEG1, and the acute neurophysiological and strength modifications (i.e., V2_RATIO_), across all participants (n = 19). Statistical significance was set at p ≤ 0.05, with p-values adjusted using the Bonferroni correction.

|  | **Grip** |  | **SICI** |  | **MEP** |  | **SICI/MEP** |  | **H80** |  | **TVR** |  |
| --- | --- | --- | --- | --- | --- | --- | --- | --- | --- | --- | --- | --- |
|  | *r* | *p* | *r* | *p* | *r* | *p* | *r* | *p* | *r* | *p* | *r* | *p* |
| **Pooled** | 0.421 | 0.432 | -0.227 | 1.000 | -0.432 | 0.564 | 0.038 | 1.000 | -0.346 | 1.000 | -0.589 | **0.048** |
| **ERSP<sub>REST</sub>** | 0.382 | 0.636 | -0.321 | 1.000 | -0.339 | 1.000 | -0.193 | 1.000 | -0.383 | 1.000 | 0.307 | 1.000 |

### Chronic effects

**Supplemental Table A.4:** Results for the chronic impact of LMV training. Repeated measures ANOVA, or Friedman test, with a single factor, Visits (V2_PRE_, V3, V4). p < 0.05 is significant.

|  | *Visits* |  |  |
| --- | --- | --- | --- |
| | $\chi^2$ | $p$ | $W$ |
| **SICI** | 0.875 | 0.646 | 0.027 |
| **MEP** | 3.125 | 0.210 | 0.098 |
| **SICI/MEP** | 0.875 | 0.646 | 0.027 |
| **H80** | 1.800 | 0.407 | 0.090 |
| **Mmax** | 2.000 | 0.368 | 0.053 |
| | $F$ | $p$ | $\eta_p^2$ |
| **RMT** | 1.978 | 0.156 | 0.117 |
| **Grip** | 1.017 | 0.352 | 0.053 |
| **M(H80)** | 1.627 | 0.234 | 0.153 |

**Supplemental Figure A.3:**
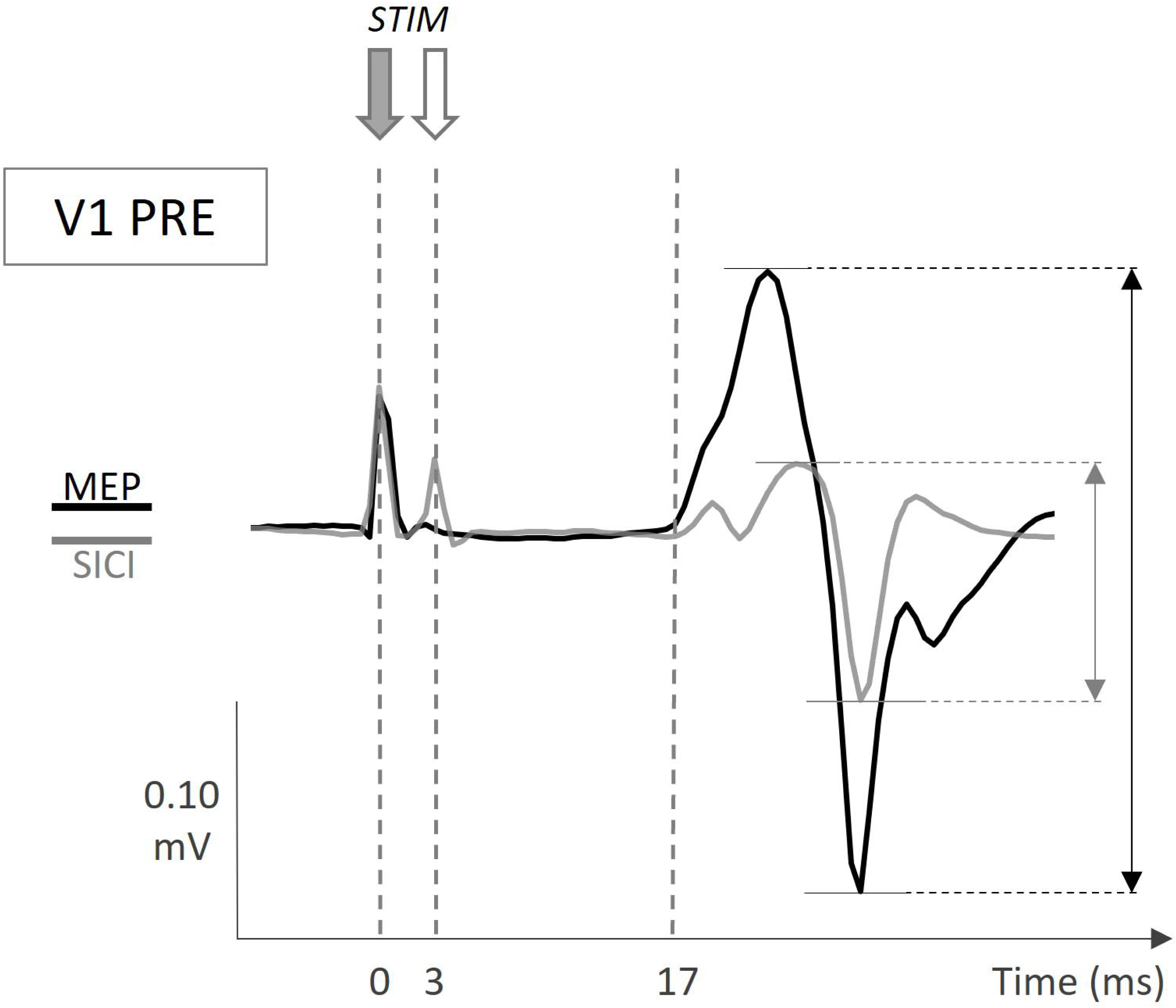
Evolution of the M(H80) in V2_PRE_ (the baseline), V3 (after 9 LMV sessions), and V4 (5 days after LMV stopped). Individual data are represented in blue and the mean of all participants is represented by the grey column. %Mmax = expressed as a percentage of maximal M-wave.

**Supplemental Table A.5:** Results of the linear Pearson’s correlation between the mean subjective pooled scores recorded during V2 and V3, or the objective measure (mean ERSP_REST_) obtained during EEG1 and EEG2, and the chronic neurophysiological and strength modifications (i.e., between V2_PRE_ and V3), across all participants (n = 19). Statistical significance was set at p ≤ 0.05, with p-values adjusted using the Bonferroni correction.

|  | Grip |  | SICI |  | MEP |  | SICI/MEP |  | H80 |  |
| --- | --- | --- | --- | --- | --- | --- | --- | --- | --- | --- |
|  | r | p | r | p | r | p | r | p | r | p |
| **Pooled** | 0.139 | 1.000 | 0.173 | 1.000 | -0.120 | 1.000 | 0.298 | 1.000 | 0.259 | 1.000 |
| **ERSP<sub>REST</sub>** | 0.108 | 1.000 | 0.053 | 1.000 | -0.033 | 1.000 | 0.056 | 1.000 | 0.259 | 1.000 |

